# Antibody response to a fourth mRNA Covid-19 vaccine boost in weak responder kidney transplant recipients

**DOI:** 10.1101/2021.09.03.21262691

**Authors:** Sophie Caillard, Olivier Thaunat, Ilies Benotmane, Christophe Masset, Gilles Blancho, on behalf of the French-speaking Society of Transplantation

## Abstract

The US FDA has recently authorized immunocompromised people to receive a third dose of mRNA Covid-19 vaccine following the two-doses regimen to further boost protection. Unfortunately, a non-negligible proportion of people treated with immunosuppressive drugs either do not respond or show only a weak response after a third boost and should, therefore, still be considered at risk of severe Covid-19. As of June 2021, we were granted the opportunity to offer a fourth vaccine dose to French solid organ transplant recipients who still showed a weak antibody response after the third dose. In this multicenter study, we demonstrate that that the protection conferred by a fourth dose is adequate for the majority of kidney transplant recipients.

---

Kidney transplant recipients (KTR) on immunosuppressive drugs have impaired immune responses to mRNA Covid-19 vaccines (1). Consequently, this population remains at high risk of severe disease during the ongoing fourth wave of the pandemic driven by the delta variant. The US FDA has recently authorized immunocompromised people to receive a third dose of vaccine following the two-dose regimen to further boost protection. French health authorities had approved the third vaccine dose on April 11, 2021 and several groups have since reported their results (2,3). Approximately 50% of patients who failed to respond after the second dose, seroconverted after a third booster dose. However, despite the fact that a three-dose scheme of vaccination allows reaching a seroconversion rate of up to ∼65% in KTR, it can still be argued that patients with low titers of anti-spike IgGs remain insufficiently protected against severe Covid-19 because they usually lack neutralizing antibodies, which is currently accepted as the best available correlate of protection. Anti-spike IgG titer above 143 BAU/mL was indeed found to correlate well with the presence of neutralizing antibodies against the wild-type virus and alpha, beta and gamma variants, while neutralization of the delta variant requires higher anti-spike IgG titers (4).

Starting from these premises, in June 2021 the French health authorities allowed offering a fourth vaccine dose to weak responder solid organ transplant recipients. A total of 92 kidney transplant recipients whose anti-spike IgG titer was between 1 and 143 BAU/mL (27 women, 65 men, median age: 58.8 years, IQR: 51-67) from three independent centers (Strasbourg, Lyon, and Nantes, France) received a fourth booster dose (BNT162b2 n=34, mRNA-1273 n=58) and had their anti-spike IgG titers measured 2 to 6 weeks thereafter. The serology assessment of IgG response was performed using the ARCHITECT IgG II Quant test (Abbott), Maglumi (Snibe) or Roche Elecsys assays. According to the test manufacturers, IgG titer>1 BAU/mL were considered positive. The study protocol complied with the tenets of the Helsinki Declaration and was approved by the Institutional Review Board (approval number: 18/21 03, Comité de Protection des Personnes Ouest IV Nantes).

There were no safety concerns with the fourth vaccine dose. The median anti-Spike IgG level increased from 16.6 (IQR: 6.5-70.1) to 146.2 BAU/mL (IQR: 28.5-243, p<0.001; Figure 1) after a median of 29 days and 54.3% of patients reached the threshold of 143 BAU/mL. Conversely, patients who remain non-responders after the third dose (< 1 BAU/ml) are unlikely to respond to a fourth boost (data not shown) and may instead benefit from pre- or post-exposure infusions of monoclonal antibodies as an alternative prophylactic option (5). In conclusion, a fourth booster vaccine dose aiming at raising the titer of anti-spike IgGs should be offered to KTR who failed to respond adequately after three doses.

**Figure 1.**
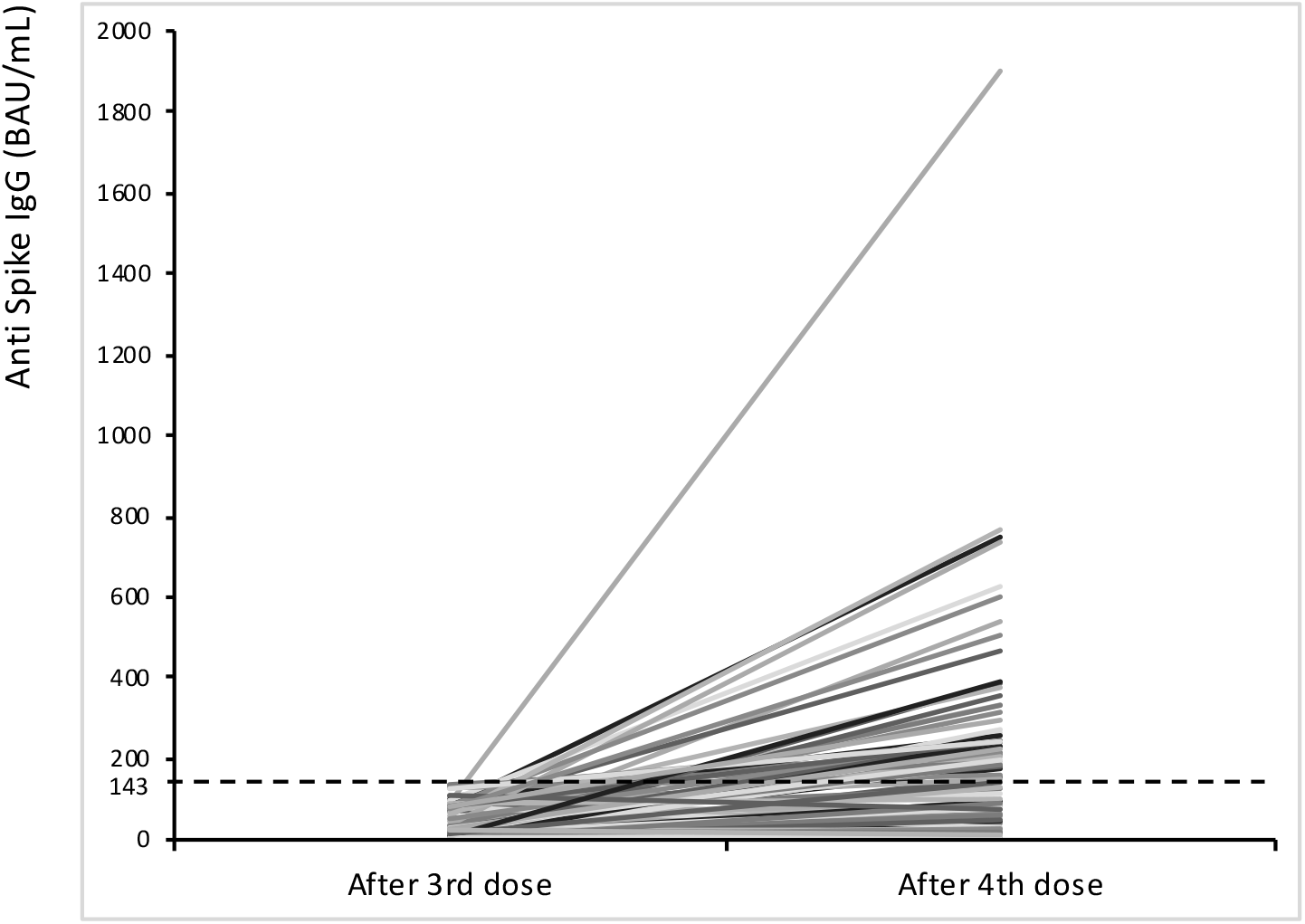
Anti-Spike IgG titers 2 to 6 weeks after the 3^rd^ and 4^th^ vaccine boost in 92 kidney transplant recipients, expressed in Binding Antibody Units (BAU), titers calibrated to the World Health Organization standard. Threshold of 143 BAU/mL is represented by the dotted line.

## Data Availability

Data are available on request

No conflict of interest to disclose

No funding

